# Experience-Based Co-Design of the Virtual Physiotherapist-led Evaluation of Referrals for spine surgery (VIPER) model of care: study protocol

**DOI:** 10.1101/2025.10.15.25338120

**Authors:** Andrew R Gamble, Giovanni E Ferreira, Christopher G Maher, David B Anderson, Joshua M Hutton, Tarcisio F de Campos, Sophie MacPherson, Christopher S Han, Mouna Sawan, Ian Harris, Sam Adie, Leanne Hassett, Christopher Legg, James Van Gelder, Mark Halliday, Monika Boogs, Rowena Charteris, Christopher Williams, Laurent Billot, Deborah Fullwood, Edward Riley-Gibson, Emma Molineux, Karen Tambree, Maria Tchan, Saurab Sharma, Joshua R Zadro

## Abstract

**Background:** Despite guidelines advising that most low back pain is best managed in primary care without surgery, many patients referred to spine surgeons have not tried effective non-surgical care. These patients can wait 1-2 years to see a surgeon and develop symptoms which become more complex and costly to manage. A promising solution is the ‘Virtual Physiotherapist-led Evaluation of Referrals for spine surgery (VIPER)’ model of care which involves a physiotherapist triaging new patient referrals suitability for a course of non-surgical care. Participants will be discharged if they no longer need to see a surgeon or referred back to see the spine surgeon if they are not responding to physiotherapist-led treatment. Evaluation of similar models in Australia have shown 2 in 3 referrals for non-urgent musculoskeletal conditions could be appropriately managed by a physiotherapist without needing to see a surgeon.

**Aims:** The aim of this study is to co-design the VIPER model of care so that it is user-centred and acceptable to patients, clinicians and key stakeholders.

**Methods:** The co-design and evaluation of the new model of care will be conducted across three phases. The first phase involved qualitative interviews to explore stakeholders’ perceptions of the proposed model of care. The second phase will involve co-design of the model of care with patients, clinicians and key stakeholders. This co-design study will use an Experience-Based Co-Design methodological approach to develop a final model of care for evaluation (third phase) in a randomised controlled trial in Australian public hospital spine surgery clinics.

## 1. Introduction

Low back pain affects 4 million Australians and creates considerable health service delivery challenges in Australia.^1^ Most guidelines advise that low back pain is best managed in primary care with advice and education to support self-management, and effective physical and psychological therapies.^2^ Despite this, referrals to spine surgeons are common. In Australia there are 45,000 lumbar spine surgeries performed each year,^3^ mostly in older adults,^4^ costing over $1 billion.^3^

Increasing referrals due to an ageing population means many spine surgery clinics struggle to assess new ‘non-urgent’ cases within 1-2 years (50% of case load).^5^ In many cases these patients have not tried effective non-surgical care, and while they wait 1-2 years to see a surgeon, they develop symptoms which are complex and costly to manage.^6^ This problem compounds in rural/remote areas where waiting times are even longer.^7^

A promising solution that optimises existing healthcare workforce capacity is a triage and management service where physiotherapists screen ‘non-urgent’ referrals to spine surgery clinics and determine their suitability for non-surgical management. Similar models have been explored in Australia.^8^ For example, a retrospective study of this model in a Victorian Hospital found that 2 in 3 referrals for a non-urgent musculoskeletal condition could be appropriately managed by a physiotherapist without needing to see a surgeon.^8^ Unfortunately, there is yet to be a high-quality clinical trial evaluating this model in Australian and globally no one has explored a virtual service to ensure equitable access.

The co-design and evaluation of the VIPER model of care will be conducted across three phases. The first phase involved qualitative interviews to explore stakeholders’ perceptions of the proposed model of care.^9^ The second phase will involve co-design of the model of care with patients, clinicians and key stakeholders. Co-design is defined as ‘a collaborative approach that actively involves service users, providers, and other stakeholders in service design, ensuring those most affected have a meaningful role in shaping decisions’.^10,11^ The third phase will be the evaluation of the VIPER model of care in a randomised trial to be conducted in Australian public hospital spine surgery clinics.

## 2. Aims

The aim of this study is to co-design the VIPER model of care so that it is user-centred and acceptable to patients, clinicians and key stakeholders (e.g., patients with low back pain, spine surgeons, physiotherapists, hospital department heads) for clinical evaluation.

## 3. Methods

This co-design study will use an Experience-Based Co-Design (EBCD) methodological approach to develop a final Virtual Physiotherapist-led Evaluation of Referrals for spine surgery (VIPER) model of care. EBCD emphasises capturing experiences, identifying priorities, and working together to design feasible improvements. The project will be conducted in collaboration with The Centre for Impact & Change and the VIPER research team between August 2025 and November 2025. The rationale for a co-design approach was the recognition that both consumers and clinicians hold critical, complementary knowledge: consumers bring lived experience of ongoing pain and recovery, while clinicians bring expertise in treatment and system constraints.

### 3.1 Virtual Physiotherapist-led Evaluation of Referrals for spine surgery (VIPER) model of care

In summary, the VIPER model of care (Figure 1) involves a senior physiotherapist triaging new referrals to either non-surgical care or surgery via a videoconference or in-person assessment, based on patient preference. Assessment will be guided by (our partner) NSW ACI’s 2024 audit indicators for spine surgery.^12^ The triaging physiotherapist will discuss all management decisions with the treating spine surgeon via video conference or email.

**Figure 1.**
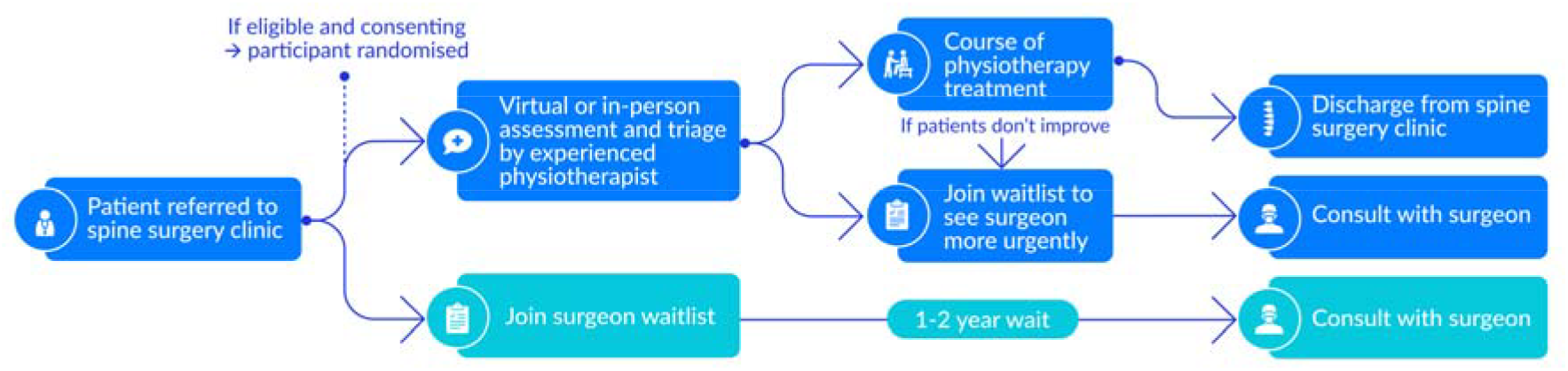
The Virtual Physiotherapist-led Evaluation of Referrals for spine surgery (VIPER) model of care

If the triaging physiotherapist deems participants suitable for non-surgical care, they will be booked for a course of in-person and/or telehealth physiotherapy (based on participant preference and to improve access in rural/remote areas). They will then receive care as typically provided in NSW Public Hospitals. These participants will be discharged if they no longer need to see a surgeon or be referred back to see the spine surgeon if they are not responding to physiotherapy treatment after 6 months (typically how long non-surgical care is recommended before considering surgery). Participants referred back to see the surgeon won’t lose their original place in the queue. If the triaging physiotherapist initially deems participants as needing to see a surgeon, they will join the wait list for the next available appointment (as per all participants in the usual care group).

### 3.2 Study Setting and Sample

Co-designers include health consumers with experience of low back pain (patients), clinicians with experience in the management of patients with low back pain (orthopaedic surgeons, neurosurgeons, physiotherapists, psychologists, and general practitioners), key stakeholders interested or involved in policy or organisations which may impact implementation (e.g. heads of physiotherapy, orthopaedic or neurosurgery departments at hospital sites in NSW, representatives from the Australian Physiotherapy Association) and the VIPER research team. Both orthopaedic surgeons and neurosurgeons commonly perform spine surgery so will be included in our sample. An external facilitator (MT) with expertise in co-design in health care settings will design the workshop processes, guide discussions, and ensure balanced participation across groups. All participants will provide written informed consent before participating the co-design workshops.

We will recruit patients via a brief advertisement on social media platforms X, LinkedIn and Facebook (Appendix 1). The advertisement will briefly describe the study and provide a link to a welcome page of the survey providing more detail, as well as the Participation Information Statement (Appendix 2) and consent form (Appendix 3). We will also recruit participants via our existing network of health professionals’ collaborators who will send an individualised email invitation (Appendix 4) to patients who may be eligible to participate, as well as an individual email invitation as an expression of interest to patients who participated in the phase 1 VIPER qualitative study. We will also use passive snowballing to recruit patients, where participants will be asked to discuss the research with others who may be interested (and eligible) to participate. These potential participants will then be invited to contact the research team to discuss their involvement if they are interested. The email invitation will briefly explain the study and contain a Participant Information Statement (Appendix 2) and consent form (Appendix 3).

We will recruit clinicians and key stakeholders through social media and our existing network of collaborators with a formal invitation email, seeking an expression of interest from individuals. A brief advertisement on social media platforms X, LinkedIn and Facebook (Appendix 1) will describe the study and provide a link to a welcome page of the survey providing more detail, as well as the Participation Information Statement (Appendix 5) and Consent Form (Appendix 3). The email to be used for recruitment is provided in Appendix 6. The email will use the blind copy (bcc:) function to ensure staff privacy (where a group email is sent) and will, without coercion or pressure, briefly describe the study, the compensation available to participants, and a telephone number and email address for further questions. We will also contact relevant professional organisations and seek their permission to advertise the study through their mailing lists or on their websites (e.g. Australian Orthopaedic Association, Spine Society of Australia, Royal Australian College of General Practitioners, Rural Doctors Association of Australia, The Australian Rheumatology Association, Australian Pain Society), and use a passive snowballing recruitment technique (as described above) by asking participating clinicians and key stakeholders to refer or forward the invitation email to their colleagues. These potential participants will then be invited to contact the researchers to discuss their involvement if they are interested. The email invitation will briefly explain the study and contain a clinician and key stakeholder Participant Information Statement (Appendix 5) and consent form (Appendix 3).

### 3.3 Co-design process

The co-design study will be guided by the EBCD methodological approach, a participant service improvement methodology that brings consumers and clinicians together to identify ‘touchpoints’ in care, and co-design solutions that improve both experience and outcomes.^13^ It is characterised by its focus on eliciting lived experience, creating empathy across different perspectives, and enabling joint decision making between health consumers and clinicians.^14^

While the classic EBCD cycle involves filmed consumer interviews, extended timelines, and multiple working groups, the VIPER study will adopt a streamlined process that retained the core methodological stages of EBCD while adapting them to project scope and resources.

Key adaptations will include the use of facilitated workshops in place of filmed interviews, and a three-workshop cycle that moved from separate elicitation of consumer and clinician perspectives to joint prioritisation and design. This approach aligns with recent adaptations of EBCD in diverse settings, including accelerated forms applied to service improvement in chronic disease management^15^ and breast and lung cancer services.^16^

### 3.3.1 Pre-design

Before we initiate the co-design workshops, we have had discussions with various health consumers and clinicians with experience in low back pain management, including orthopaedic surgeons, neurosurgeons, GPs and physiotherapists, and conducted a qualitative study (phase 1 in Figure 2) to understand stakeholder perceptions of the feasibility, acceptability and implementation of the VIPER model of care.^9^ Our preparation for co-design has involved contracting an independent facilitator (MT) and we will recruit our co-designers, develop and test co-design activities, and prepare an online platform and equipment for workshops.

**Figure 2.**
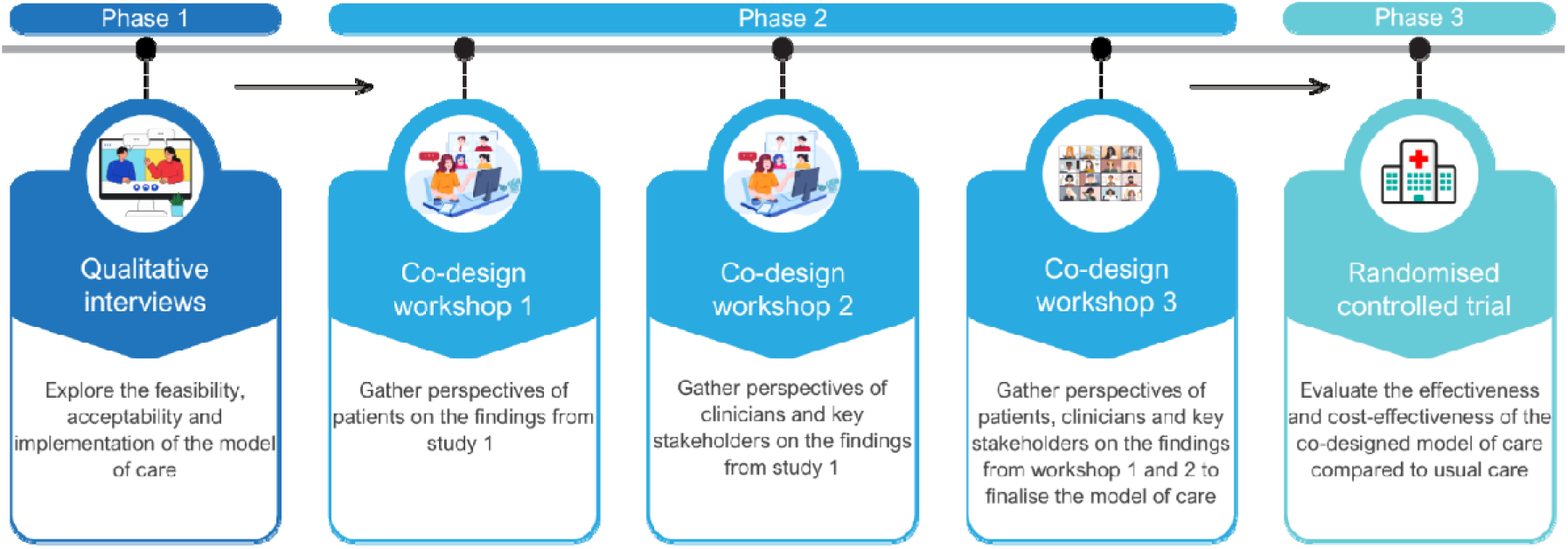
Phases of the co-design and evaluation of the Virtual Physiotherapist-led Evaluation of Referrals for spine surgery (VIPER) model of care

### 3.4 Co-design

We will conduct three workshops via Zoom (Figure 2), each lasting approximately 90–120 minutes (Workshop facilitation guide is described in Appendix 7). A design thinking approach and user-driven development process will support exploration and collaborative decision-making.^17^ Workshop activities will be tailored to the different stakeholder groups. For consumers, the nominal group technique (NGT) method will structure idea generation, ensuring equal participation, and prioritising options through group consensus.^18^ These approaches will collectively operationalise the EBCD principles of eliciting lived experience, identifying critical touchpoints, and moving from experience to solution through structured dialogue and co-creation. The purpose of each workshop is summarised in Table 1.

**Table 1:**
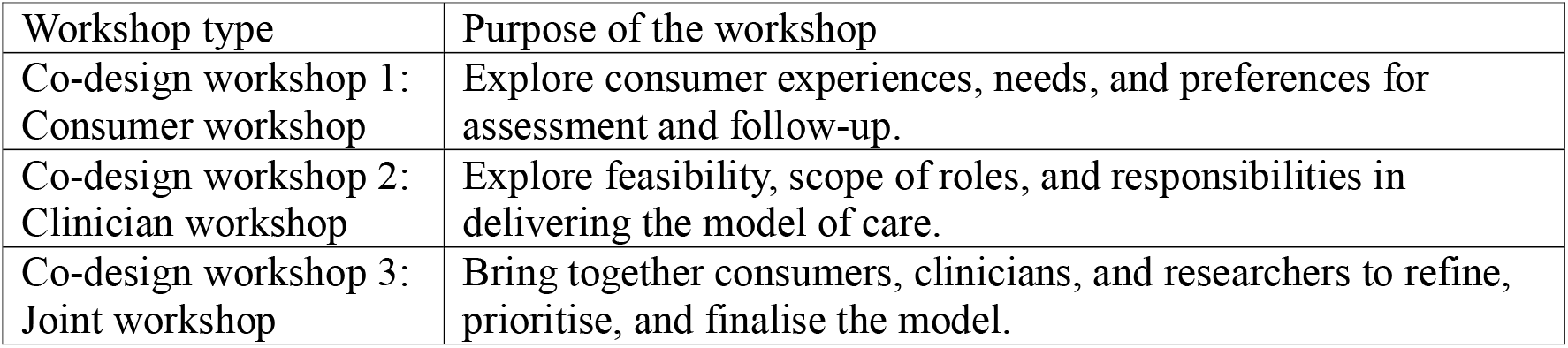
Purpose of the co-design workshops.

| Workshop type | Purpose of the workshop |
| --- | --- |
| Co-design workshop 1: Consumer workshop | Explore consumer experiences, needs, and preferences for assessment and follow-up. |
| Co-design workshop 2: Clinician workshop | Explore feasibility, scope of roles, and responsibilities in delivering the model of care. |
| Co-design workshop 3: Joint workshop | Bring together consumers, clinicians, and researchers to refine, prioritise, and finalise the model. |

Engagement strategies will include setting ground rules for respectful participation and using a range of interactive activities such as vignette testing, ranking, and consensus-building. At each stage, participants will be shown how their contributions from earlier workshops are carried forward into subsequent sessions.

Meeting minutes will be documented, meetings will be audio-recorded and transcribed, and photos will be taken of artefacts generated during the sessions.

Decision-making will combine open group discussion with structured consensus techniques. When consensus is reached, results will be immediately displayed (for example, live poll results shared on screen). Where differences remained, these will be documented and carried forward for further consideration in subsequent iterations of the model.

### 3.4 Data analysis

For data analysis, workshop discussions will be audio-recorded, transcribed, and analysed using a framework analysis approach.^19^ This method was chosen because it accommodates both structured objectives and participant-led insights, making it particularly appropriate for co-design research. An initial coding framework will be developed by the co-design workshop facilitator (MT) from the workshop objectives (e.g., telehealth assessment, care coordination role, surgeon involvement) and refined in collaboration with the VIPER research team. Additional codes added inductively as new issues emerged. Transcripts will then be coded, charted into matrices, and compared across participant groups (consumer, clinicians) and workshop types. Outputs from consensus activities (e.g., polling, prioritisation exercises) and visual artefacts (slides, draft models) will be integrated to corroborate findings and document decision-making. Rigour will be supported by triangulation of data sources, participant validation via iterative feedback, reflexive discussions between the research team and the external facilitator, ensuring findings will reflect both structured objectives and participant-led insights.

## Author Approval

All authors have seen and approved the manuscript.

## Availability of data and material

N/A as this is a protocol.

## Supplementary files

Appendix 1: Social media advertisement

Appendix 2: Participant Information Statement for patients

Appendix 3: Participant Consent Form

Appendix 4: Patient email invitation template

Appendix 5: Participant Information Statement for clinicians and key stakeholders

Appendix 6: Clinician and key stakeholder email invitation template

Appendix 7: Co-design workshop guide

