## Appendix 1 for "Experience-Based Co-Design of the Virtual Physiotherapist-led Evaluation of Referrals for spine surgery (VIPER) model of care: study protocol"

Appendix 1: Social Media Advert

**PATIENT RECRUITMENT**

**Facebook and LinkedIn advert:**

Calling all people with lived experience of low back pain!

**What?**

We are conducting a research study to understand people’s perceptions about a model of care that aims to reduce wait times to see a spine surgeon in Australian public hospitals.

**Who?**

If you have lived experience of low back pain, we invite you to participate in our important study.

**How?**

Click on the survey link below to learn more about this research [link]

Participation will involve two online workshops via Zoom (2-3 hours each)

We greatly appreciate you for supporting this important research.

**Twitter/X advert:**

Do you have lived experience of low back pain?

We want to understand your view on a model of care that aims to reduce wait times to see a spine surgeon in Australian public hospitals.

Participation will involve two online workshops via Zoom (2-3 hours each). Click on the link below learn more and participate!

[link]

**CLINICIAN RECRUITMENT**

**Facebook and LinkedIn advert:**

Calling all Australian clinicians who treat low back pain!

**What?**

We are conducting a research study to understand people’s perceptions about a model of care that aims to reduce wait times to see a spine surgeon in Australian public hospitals.

**Who?**

If you are a registered clinician in Australia who manages people with low back pain, we invite you to participate in our important study.

**How?**

Click on the survey link below to learn more about this research [link]

Participation will involve two online workshops via Zoom (2-3 hours each).

We greatly appreciate you for supporting this important research.

**Twitter/X advert:**

Are you a clinician in Australia that treats low back pain?

We want to understand your view on a model of care that aims to reduce wait times to see a spine surgeon in Australian public hospitals!

Participation will involve two online workshops via Zoom (2-3 hours each). Click on the link below learn more and participate!

[link]
