## Appendix 2 for "Experience-Based Co-Design of the Virtual Physiotherapist-led Evaluation of Referrals for spine surgery (VIPER) model of care: study protocol"

### Participant Information Statement

Dr Andrew Gamble

Faculty of Medicine and Health, School of Public Health

#### 1. What is this study about?

We are conducting a research study about a new proposed model of care is ‘Virtual Physiotherapist-led Evaluation of Referrals for spinal surgery’ (VIPER). VIPER will ensure people with low back pain who do not need surgery see a physiotherapist for effective care sooner, and those who need surgery see a surgeon sooner. VIPER involves a senior physiotherapist triaging new referrals to spine surgery clinics via videoconference, guided by (our partner) NSW ACI’s new audit indicators for spine surgery. Participants suitable for non-surgical care will be booked for an in-person or telehealth physiotherapy appointment (based on preference and to improve access in rural/remote areas) and receive care as typically provided in NSW Public Hospitals (e.g., advice to support self-management, exercise therapy). These participants will be discharged if they no longer need to see a surgeon.

You are invited to take part in a research study to help codesign the proposed VIPER model of care to inform its adaptation in a trial.

Taking part in this study is voluntary. Please read this sheet carefully and ask questions about anything you don’t understand or want to know more about.

#### 2. Who is running this study?

The study is being carried out by the following researchers:

- Dr Andrew Gamble, Research Fellow, The University of Sydney
- Dr Joshua Zadro, Research Fellow, The University of Sydney
- Dr Giovanni Ferreira, Research Fellow, The University of Sydney
- Prof Chris Maher, Research Professor, The University of Sydney
- Dr David Anderson, Research Fellow, The University of Sydney
- Dr Tarcisio Folly De Campos, Research fellow, The University of Sydney
- Dr Chris Han, Research Assistant, The University of Sydney
- Ms Sophie Macpherson, Research Assistant, The University of Sydney

This study is being funded by a HCF Health Services Research Grant. The study is sponsored by The University of Sydney. There are no potential or actual conflicts of interest or financial benefits to the researchers, sponsor or institutions from this research.

#### 3. Who can take part in the study?

This study is recruiting people with a lived experience of low back pain. You are eligible if you have currently, or previously experienced low back pain and living in Australia.

This Participant Information Statement tells you about the research study. Knowing what is involved will help you decide if you want to take part in the research. Please read this sheet carefully and ask questions about anything that you don’t understand or want to know more about. Participation in this research study is voluntary.

By giving your consent to take part in this study you are telling us that you:

- Understand what you have read.
- Agree to take part in the research study as outlined below.
- Agree to the use of your personal information as described.

You will be given a copy of this Participant Information Statement to keep.

#### 4. What will the study involve for me?

If you agree to participate in this study, you will be asked to complete the consent form. You may then be asked to attend 2 group-based workshops, but this is not guaranteed. We aim to recruit people with diverse characteristics and opinions. The group-based workshops will be conducted virtually via Zoom; they will be between 2-3 hours in length each workshop and will be recorded. The aim of the workshops is to draw on your experience to provide feedback on the proposed model of care before it is implemented in the VIPER trial. The workshops will be facilitated by a co-design expert from the company – The Centre for Impact & Change. The facilitator will guide the workshops based on the responses and a series of activities to prompt responses.

Workshop 1 Objective: Gather patient perspectives on the qualitative study findings and proposed model of care.

Workshop 2 Objective: Bring together patient, clinicians’ and key stakeholders’ feedback to finalize the model of care.

All aspects of the study, including results, will be strictly confidential and only the study investigators will have access to the data. The data from this study may be used again in the future, for further research purposes within the research team. All your identifying data will remain confidential. Study materials will be stored on The University of Sydney’s Research Data Store (RDS) system. Data will be kept in perpetuity, but future projects that aim to use the data will seek ethics approval before using such data. A report of the study may be presented at a conference or in a scientific journal, but individual participants will not be identifiable in such a report.

#### 5. Can I withdraw once I have started?

Participation in this study is entirely voluntary. You are not obliged to participate. If you do participate, you can withdraw at any time without having to give any reason and without suffering any penalty. If you take part in the codesign workshops, you are free to stop at any stage or to refuse to answer any of the questions. However, since it is a group discussion, it may not be possible to withdraw your individual comments from our records once the workshop has started and any information that we have already collected will be kept and will be included in the study results. Whatever your decision, it will not affect your relationship with The University of Sydney and University of New South Wales.

#### 6. Are there any risks or costs?

Aside from giving up your time, we do not expect that there will be any risks or costs associated with taking part in this study.

#### 7. Are there any benefits?

By participating you will be contributing to important research that helps us finalise the VIPER model of care for people with persistent pain after knee replacement surgery. You will also be compensated for your time at an hourly rate of $50 per hour post attendance of the two workshops.

#### 8. What will happen to information that is collected?

By providing your consent, you are agreeing to us collecting personal information about you for the purposes of this research study. Your information will only be used for the purposes outlined in this Participant Information Statement, unless you consent otherwise. Your information will be stored on University of Sydney servers.

Your identity/information will be kept strictly confidential, except as required by law. Study findings may be published, but you will not be individually identifiable in these publications.

We will keep the information we collect for this study, and we may use it in future projects. By providing your consent you are allowing us to use your information in future projects and share it locally and internationally with other research collaborators as needed. We don’t know at this stage what these other projects will involve. We will seek ethical approval before using the information in these future projects.

We will use Microsoft Word’s or Otter ai’s transcription feature to transcribe workshops. This will involve sharing your information with Microsoft or Otter ai. We will not share this information with anyone else without your consent unless we are required to do so by law. Microsoft word is owned by Microsoft and located in Redmond, Washington, US and Otter.ai, Inc. is a transcription software company based in Mountain View, California.

We will store this information and dispose of it securely following the University’s Recordkeeping Policy. For more details about how your information will be handled please see the University’s [privacy webpage](https://www.sydney.edu.au/about-us/governance-and-structure/privacy-and-university-information/privacy-at-the-university.html).

Sharing research data is important for advancing knowledge and innovation. A de-identified set of the data collected in this study may be made available for use in future research.

#### 9. Will I be told the results of the study?

You have a right to receive feedback about the overall results of this study. You can tell us that you wish to receive feedback by selecting ‘yes’ to feedback on the written consent form. This feedback will be in the form of a one-page lay summary of the results. You will receive this feedback after the study is finished. Feedback regarding personal results will not be available.

#### 10. What if I would like more information?

When you have read this information, please store it in a safe place. The study researchers will be available to discuss the study with you and answer any questions you may have. If you would like to know more at any stage, please feel free to contact the VIPER research study team on (02) 8627 6691 or.

#### 11. What if I have a complaint or any concerns?

The ethical aspects of this study have been approved by the Human Research Ethics Committee (HREC) of The University of Sydney [ethics reference: 2025/HE…] according to the National Statement on Ethical Conduct in Human Research. This statement has been developed to protect people who agree to take part in research studies.

If you are concerned about the way this study is being conducted or wish to make a complaint to someone independent from the study, please contact the university using the details outlined below. Please quote the study title and protocol number.

The Manager, Ethics Administration, University of Sydney:

-

***This information sheet is for you to keep***
