## Appendix 3 for "Experience-Based Co-Design of the Virtual Physiotherapist-led Evaluation of Referrals for spine surgery (VIPER) model of care: study protocol"

### Participant Consent Form

Dr Andrew Gamble

Faculty of Medicine and Health, School of Public Health

| **Participant Name** |
| --- |

I agree to take part in this research study. In giving my consent, I confirm that that:

- The details of my involvement have been explained to me, and I have been provided with a written Participant Information Statement to keep.
- I understand the purpose of the study is to investigate the proposed Virtual Physiotherapist-led Evaluation of Referrals for spine surgery (VIPER) model of care.
- I acknowledge that the risks and benefits of participating in this study have been explained to me to my satisfaction.
- I understand that in this study I will be required to attend two virtual group-based workshops.
- I understand that participation involves recording using Zoom software of the group workshops, the recording will be deleted after the transcription is finalized.
- I understand that if I provide consent my information may be used in future research purposes within the research team and be presented a conference of in a scientific journal.
- I understand that being in this study is completely voluntary.
- I am assured that my decision to participate will not have any impact on my relationship with the research team or the University of Sydney.
- I understand that I am free to withdraw from this study at any time and that I can choose to withdraw any information I have already provided (unless the data has already been de-identified or published).
- I have been informed that the confidentiality of the information I provide will be protected and will only be used for purposes that I have agreed to. I understand that information identifying me will only be told to others with my permission, except as required by law.
- I understand that the results of this study may be published, and that publications will not contain my name or any identifiable information about me.
- I confirm the following:

| **I consent to recordings of the group workshops** | Yes **☐** No **☐** |
| --- | --- |
| **I consent to** **being contacted for future studies** | Yes **☐** No **☐** |
| **I would like feedback on the overall results of this study** | Yes **☐** No **☐** |

If you answered **yes** to receiving feedback or being contacted in future, please provide your preferred contact details (email/telephone/postal address):

- I understand that after I sign and return this consent form it will be retained by the researcher, and that I may request a copy at any time.

| **Participant Name** |
| --- |
| **Signature** |
| **Date** |
