## Appendix 4 for "Experience-Based Co-Design of the Virtual Physiotherapist-led Evaluation of Referrals for spine surgery (VIPER) model of care: study protocol"

**Example email to patient representatives**

**SUBJECT LINE:** Recruiting patient consumers to help codesign a care model for the VIPER trial

Dear [individual],

Researchers at the University of Sydney and University of New South Wales are developing a model of care for patients with low back pain referred for surgery. You are eligible if you have currently, or previously experienced low back pain.

They are interested in eligible patients’ to participate in the project, drawing on your experience to provide feedback on the proposed model of care before it is implemented in the trial, whether it is positive or negative.

The co-design process includes two interactive, virtual group-based workshops where you will collaborate with researchers and other healthcare professionals to identify key priorities and contribute to the model’s development.

📅 Workshop 1: [TBC]

📅 Workshop 2: [TBC]

📍 **Location:** Virtual via Zoom Meeting

If you would like to participate, please review the attached participant information statement and complete the attached consent form and return via email to.

Thank you very much for your time.

Kind regards,
[Firstname Lastname]
