## Appendix 6 for "Experience-Based Co-Design of the Virtual Physiotherapist-led Evaluation of Referrals for spine surgery (VIPER) model of care: study protocol"

**Example email to clinicians and key stakeholder representatives**

**SUBJECT LINE:** Recruiting clinicians and key stakeholders to help codesign a model of care for the VIPER trial

Dear [individual],

Researchers at the University of Sydney and University of New South Wales are developing a model of care for patients with low back pain referred for surgery. Clinicians are eligible if you have managed at least 5 patients with low back pain within the last 12 months. Key stakeholders are eligible if you are interested or involved in policy or organisations which may impact the implementation of the VIPER model of care.

They are interested in eligible clinicians and key stakeholders to participate in the project, drawing on your experience to provide feedback on the proposed model of care before it is implemented in the trial, whether it is positive or negative.

The co-design process includes two interactive, virtual group-based workshops where you will collaborate with researchers and other clinicians or key stakeholders to identify key priorities and contribute to the model’s development.

📅 Workshop 1: [TBC]

📅 Workshop 2: [TBC]

📍 **Location:** Virtual via Zoom Meeting

If you would like to participate, please review the attached participant information statement and complete the attached consent form and return via email to.

Thank you very much for your time.

Kind regards,
[Firstname Lastname]
