## Appendix 7 for "Experience-Based Co-Design of the Virtual Physiotherapist-led Evaluation of Referrals for spine surgery (VIPER) model of care: study protocol"

**WORKSHOP GUIDE**

*Note: The topics below will serve as an outline to guide the workshops.*

**Introduction**

Introduce yourself to the workshop group.

Thank you for agreeing to participate in today’s workshop. This workshop is voluntary and at any point of time you are free to withdraw. The workshop can take a maximum of 2-3 hours of your time today.

The purpose of this workshop is to codesign the Virtual Physiotherapist-led Evaluation of Referrals for spine surgery (VIPER) model of care before it is implemented in a trial.

1. Thank participants for their time. Check that it is OK to audio record the workshop. If yes, indicate when pressing record. Advise that:
a. They can stop at any time, and continue later or not, whatever suits best.
b. All responses will be anonymous and confidential.
2. Any questions before getting started?

Note for Facilitator: Follow-up prompts will be used to explore responses. For example, “What makes you say that?” or “Could you elaborate further?” This will allow an in-depth understanding of topics and responses. As indicated following each question, more specific prompts will be used to ensure complete coverage of topics raised, ensuring that workshops are in-depth and provide relevant data.

**Informant workshop questions**

Listed below are a series of discussion prompts and activities that may be asked to participants.

a) **WORKSHOP 1: Patient Representatives Workshop**

**Objective:** Gather patient perspectives on the qualitative study findings and proposed model of care.

**Structure:**

**Introduction (20 mins):** Present study results and explain the purpose of the workshop.

- Welcome participants and introduce facilitators.
- Explain the purpose of the workshop and how their input will shape the model of care.
- Present key findings from the qualitative study.
- Provide a brief overview of the proposed model of care.

**Discussion Group (40 mins):**

- **Reflections on Patient and Clinician Feedback from the qualitative study:**
  - What are your thoughts on the concerns raised by the patient interviews?
  - What are your thoughts on the concerns raised by the clinician interviews?
- **Model of Care Review:**
  - Do you see your experiences reflected in the proposed model?
  - Are there any missing elements that should be included?
  - Are there any aspects that seem unnecessary or impractical?

**Group Feedback Session (40 mins):** Consolidate key themes from the discussion.

**Wrap-Up & Next Steps (20 mins):** Summarize insights and explain the next steps.

- Explain how their feedback will be incorporated into the next phase.
- Invite any final comments or questions.

b) **WORKSHOP 2: Clinician and Key Stakeholder Representatives Workshop**

**Objective:** Gather Clinician and key stakeholder perspectives on the qualitative study findings and proposed model of care.

**Structure:**

**Introduction (20 mins):** Recap patient feedback and qualitative study outcomes.

- Welcome clinicians and key stakeholders and introduce facilitators.
- Present key findings from the qualitative study.
- Provide an overview of the proposed model and its key components.

**Discussion Group (40 mins):**

- **Reflections on Patient and Clinician Feedback from the qualitative study:**
  - What are your thoughts on the concerns raised by the patient interviews?
  - What are your thoughts on the concerns raised by the clinician interviews?
  - How do these findings compare with your clinical experience?
- **Model of Care Review:**
  - What aspects of the model align with best clinical practices?
  - Are there any components that might be difficult to implement?
  - What additional resources or training would be needed?
  - What would make this model more practical in clinical settings?

**Group Feedback Session (40 mins):** Consolidate key themes from the discussion.

**Wrap-Up & Next Steps (20 mins):**

- Explain how their feedback will be incorporated into the next phase.
- Invite any final comments or questions.

c) **WORKSHOP 3: Final Consensus Workshop (Patients, Clinicians and Key Stakeholders)**

**Objective:** Bring together patient, clinicians and key stakeholder feedback to finalize the model of care.

**Structure:**

**Recap (30 mins):**

- Summarize key takeaways from Workshops 1 & 2.
- Present an updated version of the model based on prior feedback.

**Collaborative Discussion (60 mins):**

- **Final Model Review:**
  - Does the revised model address the key concerns raised in previous workshops?
  - Are there any remaining gaps?
  - What aspects of the model are most important to keep?
  - How can we ensure the model is both patient-centred and clinically feasible?

**Consensus-Building Exercise (60 mins):**

- Prioritize final changes using dot voting or a structured decision-making tool.
- Discuss the results and finalize key components.

**Conclusion (30 mins):**

- Confirm final agreements on the model of care.
- Thank participants and outline next steps – finalizing the model report and commencement of VIPER trial.
